# A comparison of therapies using Eyetronix Flicker Glass and standard adhesive patches in children with anisometropic amblyopia: A randomized controlled trial

**DOI:** 10.1101/2020.07.20.20157552

**Authors:** Seung Hyun Min, Shijia Chen, Jinling Xu, Bingzhen Chen, Hui Chen, Yuwen Wang, Jiawei Zhou, Xudong Yu

**Affiliations:** McGill Vision Research, McGill University; Department of Ophthalmology, Wenzhou Medical University

**Keywords:** Amblyopia, Randomized clinical trial, Eyetronix Flicker Glass, Monocular occlusion, Children

## Abstract

**Purpose:** Recently, Eyetronix Flicker Glass (EFG) was introduced as a novel treatment for amblyopia. We conducted a randomized clinical trial to compare the efficacy of the therapies using the Eyetronix Flicker Glass and standard adhesive patches.

**Methods:** We tested 31 children (aged 4-13 years) with anisometropic amblyopia. They were assigned into one of the two treatment groups and were treated for 12 weeks. The first group was treated with the Eyetronix Flicker Glass for one hour per day (Flicker Group), and the latter with standard patches (Patching Group) for two hours per day. We considered best-corrected visual acuity (BVCA) of the amblyopic eye after 12 weeks of treatment as the primary outcome. Secondary outcomes were BVCA of the amblyopic at other timepoints of measurement (before the treatment, 3 and 6 weeks) as well as contrast sensitivity of the amblyopic eye, stereopsis and fusion range at all timepoints.

**Results:** Visual acuity in the amblyopic eye was significantly improved in both treatment groups at week 12 relative to baseline. The magnitude of improvement was similar. Contrast sensitivity of the amblyopic eye at 3, 6 and 12 cpd was large but not so at 18 cpd for both groups. Stereopsis did not improvement in both groups. However, fusion range significantly improved in the Flicker Group at week 12 relative to baseline.

**Conclusion:** After 12 weeks, EFG therapy improves amblyopic eye’s visual acuity, its contrast sensitivity and fusion range of the patients. EFG therapy improves both monocular and binocular functions.

**Clinical trial registry number:** ChiCTR2000034436 from the Chinese Clinical Trial Registry

## 1 Introduction

Amblyopia is a neurodevelopmental disorder that has numerous visual consequences such as poor visual acuity, contrast sensitivity and stereopsis [1, 2, 3, 4, 5, 6]. It arises from abnormal visual experience during the critical period when the brain function is most susceptible to changes in visual input [7, 8]. Hence, rewiring of the visual cortex and, hence, attaining normal visual experience for the visually impaired must take place before the critical period terminates. For the past 250 years, physicians have deprived the normal eye of amblyopic children for two to six hours for over the course of months to years [9]. The traditional patching therapy improves the visual acuity of the amblyopic eye by more than 2 lines of logMAR in over 50% of cases [**?**, 10]. However, it has been less than ideal for several reasons. First, it discomforts the children as it enforces them to only see through the amblyopic eye [11, 12]. Second, it has a low compliance rate due to the discomfort [13]. Moreover, it has been associated with adverse psychosocial factors such as emotional distress [14]. Furthermore, the improved visual acuity of the amblyopic eye does not ensure an improvement in binocular vision [15, 16], which is more important for everyday visual experience and self-perception [17]. Traditional patching therapy has also been prescribed with pharmacological intervention, such as the injection of atropine into the fellow eye. However, the possibility for the complication from its toxicity is not negligible [18]. Therefore, for these reasons that illustrate the fact that the traditional patching therapy is not flawless, an alternative treatment for amblyopia that can bring about both long-lasting monocular and binocular visual improvements is needed.

Recently, liquid crystal glasses (LCG) have been introduced as a device to include in an alternative therapy to standard patching [19]. They have the structure of typical glasses, but their lenses are built with liquid crystal cells that enable the lenses to be either transparent or opaque [20]. The alternation between the transparent and opaque states can be extremely rapid and appears as flicker. As is the case in the standard patching therapy, a therapy that incorporates the LCG only deprives the normal eye and, hence, can be categorized as a monocular therapy. Therefore, the lens in front of the normal eye flickers. However, unlike traditional patching which entirely deprives the normal eye throughout the treatment period, LCG therapy still allows the children to see through both eyes because the lens in front of the normal eye alternates between being opaque and transparent. LCG therapy shows promise. For instance, studies have shown that it improves the visual acuity of the amblyopic eye (i.e., monocular visual function), compliance and satisfaction of patients [19, 21, 22].

However, amblyopia has been recently understood as a complex and multifaceted disorder as it is characterized by not just poor monocular visual functions of the amblyopic eye but also imbalanced binocular interactions between the normal and amblyopic eyes [23, 24, 25, 26, 27, 28]. In normal observers, the two eyes suppress each other during normal viewing in a balanced fashion. However, in amblyopes, the suppression of the amblyopic eye on the fellow eye is slightly weaker than that of the fellow eye on the amblyopic eye [29, 28]. This results in a net imbalance resulting in total dominance by the fellow eye. Therefore, an important step to treat amblyopia based on this abnormal binocular interaction is to redress this imbalance between the eyes. The disrupted binocular balance in amblyopia might be the reason why the traditional patching therapy, which only manipulate the visual input through one eye (i.e., normal eye), can be inadequate for a full recovery [15, 16]. Conversely, a binocular treatment, which manipulates input in both eyes, could be more beneficial. Researchers have already explored this possibility. For example, Schor, Terrell and Peterson (1976) demonstrated that flicker that alternately occurs in lenses for both eyes (i.e., alternate flicker) at about 2 and 7 Hz reduces an enhanced masking by the fellow eye on the amblyopic eye [30]. In other words, the disrupted binocular interaction became more balanced in amblyopic patients, and the visual acuity of the amblyopic eye improved. This study shows that binocular treatment strategy can benefit amblyopic patients.

Based on the premise that alteration of binocular input benefits binocular functions, Eyetronix® have introduced Eyetronix Flicker Glass (EFG). In a therapy that incorporates EFG, the lenses in front of the amblyopic and normal eyes alternate between transparent and opaque states; this is similar to alternate flicker as seen in Schor et al. (1976) [30]. Hence, EFG therapy deprives inputs from both eyes in an alternate fashion unlike the LCG therapy which deprives the normal eye only [22, 31]. If it is true that binocular therapy can improve binocular functions in amblyopia, as it was reported by Schor et al. (1976), the EFG therapy might benefit binocular functions such as stereopsis and fusion. Indeed, a recent study shows that EFG therapy for 12 weeks improves both the visual acuity of the amblyopic eye and stereopsis in 23 patients with anisometropia amblyopia [32]. Furthermore, a subset of the patients showed a sustained improvement in visual acuity and stereopsis for 12 weeks after the treatment. However, although Vera-Diaz et al. observed a noticeable improvement in visual acuity and stereopsis using the EFG therapy, they did not compare the efficacy with that of the standard patching therapy [32].

In this study, we performed a randomized controlled trial for the first time to compare the efficacy of traditional patching and EFG therapies in amblyopic children. We measured monocular (amblyopic eye’s visual acuity and contrast sensitivity) and binocular visual functions (stereopsis and fusion range) before and during the treatment for over 12 weeks. Throughout this study, we regard best-corrected visual acuity measured after 12 weeks of treatment as primary outcome.

## 2 Materials and Methods

This randomized clinical trial is listed in the Chinese Clinical Trial Registry (http://www.chictr.org.cn) with the identifier ChiCTR2000034436. The Full trial can be accessed from the Chinese Clinical Trial Registry’s website: http://www.chictr.org.cn/showproj.aspx?proj=56030. The research protocol and the informed consent forms were reviewed and approved by the ethics committee of the Affiliated Eye hospital at Wenzhou Medical University (2016-18-Q-11). All data were collected at the Affiliated Eye Hospital of Wenzhou Medical University.

### 2.1 Eyetronix Flicker Glass

Eyetronix Flicker Glass (http://eyetronix.com) utilizes liquid crystal technology. Liquid crystal has properties of liquid and a solid crystal. Its liquid properties enable the molecules within to readily rotate and move and make liquid crystal an excellent conductor of light.

Each lens of the EFG contains liquid crystal shutter cells [20], which are suspended in gel-like liquid (light blue space in Figure 1A). The gel-like liquid is sandwiched between one thin glass plate on one side and another plate on the other side (thin green structures in Figure 1A). On the outer side of each thin glass plates lies a polarizer film that controls the transmission of light (striped or dotted structure in Figure 1A). Light transmission through the lenses depends on the polarizer film. Since there are two glass plates in each lens, there are two polarizer films per lens. These two polarizer films are at perpendicular orientation to one another (90°). The first polarizer only allows the horizontally oriented light to pass, and the second polarizer only the vertically oriented light because the orientations of the polarizers are perpendicular to one another (the striped pattern represents a perpendicular orientation to the dotted pattern in Figure 1A). Therefore, the light transmitted by the first polarizer will not be transmitted by the second polarizer in the lens of the EFG is in its natural state. When no external energy is applied, the lens is therefore opaque, and no light gets through (Closed/Occlusion state in Figure 1A).

**Figure 1:**
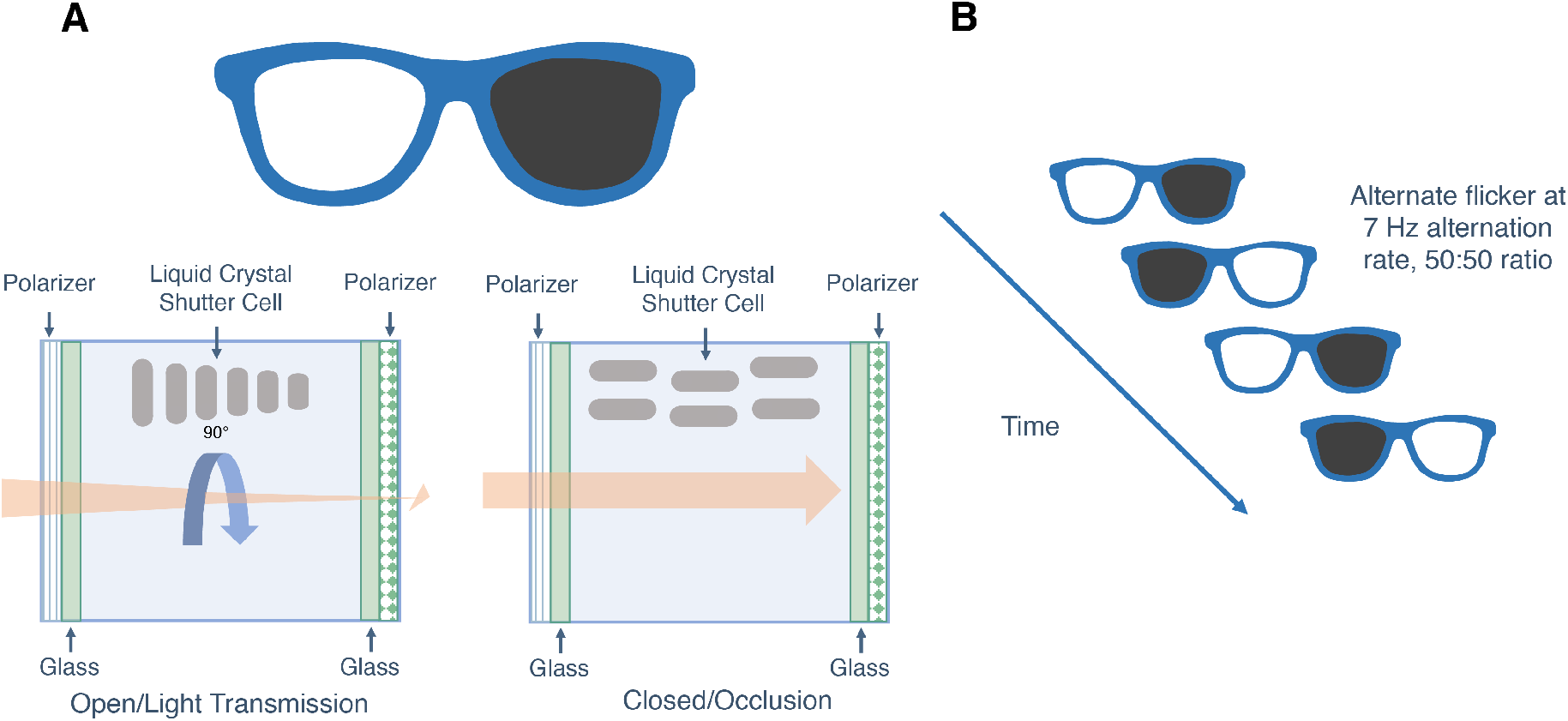
A diagrammatic representation of the Eyetronix Flicker Glass. A) The lens of the EFG can assume either a transparent or opaque state depending on whether the lens allows light transmission or not. When electric field is present, the lens assumes an ‘open’ state and enable light transmission. This will make the lens transparent. When electric field is absent, the lens assumes an ‘closed’ state and obstruct light transmission. This will make the lens opaque. B) An illustration of alternate flicker of the EFG. We programmed the EFG so that it would perform alternate flicker at 7 Hz alternation rate, 50:50 ratio between the lens in front of the amblyopic eye and that in front of the normal eye. Therefore, 1-hour treatment with the EFG would be equivalent to 30 minutes deprivation of the normal eye.

However, by applying external energy, one can make the lens to be transparent (Open/Light transmission state in Figure 1A). When an electric field is present, the formerly perpendicular orientations of the polarizer films become parallel and aligned because the liquid crystal shutter cells (shown in grey structures in Figure 1A), which are suspended in gel-like liquid, will change their position. As a result, the light will be able to be transmitted through the lens. In other words, by applying electric field, one can control make the formerly opaque liquid crystal lens to become transparent within milliseconds (see Figure 1B).

EFG and Liquid Crystal Glasses (LCG) both utilize the liquid crystal technology and share the goal of replacing the standard patching therapy. However, they are slightly different. The LCG therapy only deprives the normal eye by only allowing the lens in front of said eye to shift between its transparent and opaque state. On the other hand, the EFG therapy occludes both the normal and amblyopic eyes in an alternate fashion (see Figure 1B) at a specific ratio and temporal frequency.

### 2.2 Patients

32 children (aged 4-13) with untreated, moderate to severe unilateral and anisometropic amblyopia participated in our study. One of them received another treatment during the study and had to be excluded, so 31 children completed the study. The clinical details of the patients are provided in Table 1. The sample size was determined based on the previous studies of EFG and LCG [22, 32]. We obtained informed consent from their parent or legal guardians. Their guardians volunteered to participate in the study and accepted the random assignment of each patient to either Group 1 (traditional patching therapy) or Group 2 (therapy with the EFG).

**Table 1:** Baseline characteristics of the patients.

|  | Flicker Group |  | Patching Group |  |
| --- | --- | --- | --- | --- |
|  | N | % | N | % |
| **Gender** |  |  |  |  |
| Female | 6 | 40% | 8 | 50% |
| **Age at enrollment (years)** |  |  |  |  |
| 4 to <6 | 9 | 60% | 6 | 38% |
| 6 to <8 | 5 | 33% | 5 | 31% |
| 8 to 13 | 1 | 7% | 5 | 31% |
| Mean ± SE | 5.27 ± 0.28 | | 6.38 ± 0.61 | |
| **Distance visual acuity of amblyopic eye (logMAR)** |  |  |  |  |
| 0.15 to <0.2 | 1 | 7% | 1 | 6% |
| 0.2 to <0.3 | 3 | 20% | 3 | 19% |
| 0.3 to <0.4 | 3 | 20% | 5 | 31% |
| 0.4 to <0.5 | 3 | 20% | 2 | 13% |
| 0.5 to <0.6 | 3 | 20% | 2 | 13% |
| 0.6 to <0.7 | 1 | 7% | 0 | 0% |
| 0.7 | 1 | 7% | 3 | 19% |
| Mean ± SE | 0.39 ± 0.04 | | 0.39 ± 0.05 | |
| **Distance visual acuity of fellow eye (logMAR)** |  |  |  |  |
| Mean ± SE | 0.03 ± 0.01 | | 0.01 ± 0.02 | |
| **Interocular acuity difference (logMAR)** |  |  |  |  |
| Mean ± SE | 0.37 ± 0.05 | | 0.38 ± 0.04 | |
| **Baseline stereoacuity (arc seconds)** |  |  |  |  |
| Nil (converted to 4500) | 4 | 27% | 6 | 40% |
| 800 | 0 | 0% | 1 | 7% |
| 400 | 3 | 20% | 2 | 13% |
| 200 | 2 | 13% | 0 | 0% |
| 140 | 0 | 0% | 2 | 13% |
| 100 | 1 | 7% | 0 | 0% |
| 80 | 0 | 0% | 3 | 20% |
| 60 | 1 | 7% | 1 | 7% |
| 50 | 2 | 13% | 0 | 0% |
| 40 | 2 | 13% | 1 | 7% |
| Mean ± SE (arc seconds) | 1329.33 ± 512.11 | | 1826.25 ± 536.81 | |
| Mean ± SE (log arc seconds) | 2.49 ± 0.21 | | 2.71 ± 0.21 | |
| **Amblyopic eye's spherical equivalent (diopters)** |  |  |  |  |
| Mean ± SE | 3.95 ± 0.67 | | 4.05 ± 0.45 | |
| **Fellow eye's spherical equivalent (diopters)** |  |  |  |  |
| Mean ± SE | 0.85 ± 0.39 | | 0.77 ± 0.36 | |

logMAR = logarithm of the minimum angle of resolution; SE = standard error

#### 2.2 Patients

Eligibility inclusion criteria: 1) Age range of 4 to 13 years. 2) Definite genetic diagnosis of anisometropic amblyopia, best-corrected visual acuity (BCVA) of the amblyopic eye equal to or worse than 0.3 (logMAR) in 3–5 years age range and 0.15 (logMAR) in children over 6 years old, at least a two-line BVCA difference between the two eyes (according to the diagnostic criteria for amblyopia established by the Ophthalmology branch of the Chinese Medical Association in 2011). 3) Myopia of no more than −6.00 diopters (D), hypermetropia of no more than +9.00 D, astigmatism of less than 3.00 D, anisometropia of at least 1.50 D spherical equivalent or at least 1.00 D cylindrical equivalent. 4) Strabismus of no more than 20 prism diopters (Δ) according to prism and alternate cover test (PACT). 5) Better than a 0.7 logMAR BVCA of the amblyopic eye.

Eligibility exclusion criteria: 1) Ocular diseases, such as ptosis, refractive media opacity, fundus disease and optic neuropathy. 2) History of interocular or refractive surgery that affects vision. 3) History of treatment for amblyopia in the last three months before screening (except spectacle frames). 4) Photosensitivity epilepsy. 5) Confirmed or suspected conjunctivitis. 6) Allergy or intolerance to the test equipment or patch. 7) History of pharmacological intervention that may affect vision such as atropine. 8) Participation in another clinical study/trial within one month before the enrollment of the study.

### 2.3 Treatment procedure

After confirming for the eligibility of each patient, we randomly assigned all participants to one of the two treatment groups using a random number generator so that the group assignment was balanced for both groups (see Figure 2). EFG therapy intervention group (Flicker Group): In this treatment group, patients were instructed to wear the EFG rather than a standard adhesive patch. Schor et al. employed a rate for alternate flicker at 7 Hz [30]. Likewise, we used parameters of 7Hz and 50% duty cycle in this study. At 7 Hz, the lens assumes each state of transparent (open) and opaque (closed) state for 0.71+ seconds. At a duty cycle of 50% (50:50 ratio), EFG deprives the normal eye 50% of the time throughout the treatment. Therefore, 1-hour treatment with the EFG would be equivalent to 30 minutes deprivation of said eye (50%). The patients were asked to wear the EFG for one hour per day, 7 days per week throughout the 12-week treatment period. The EFG automatically recorded the duration and time in which the patients wore them.

**Figure 2:**
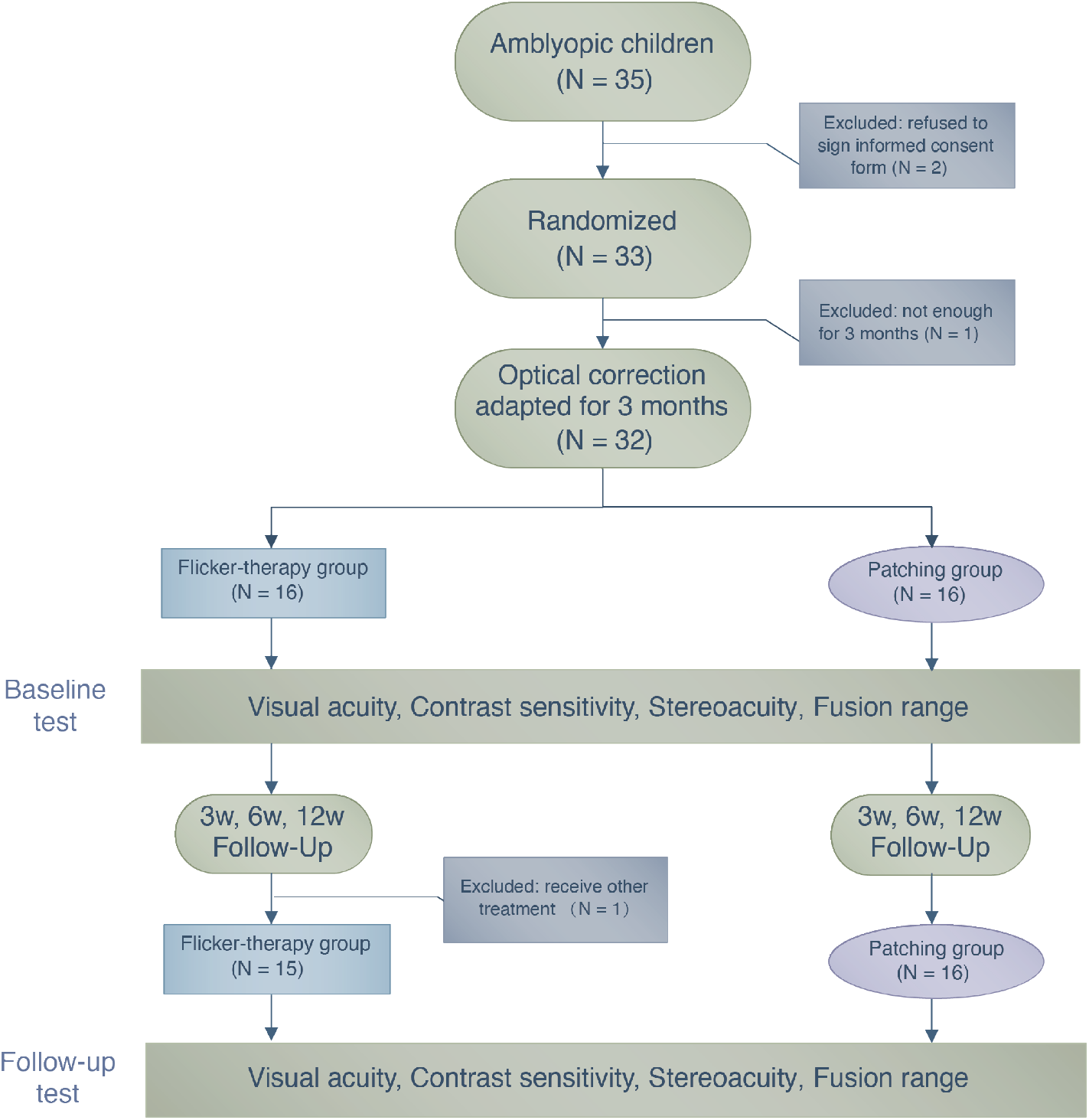
A flow chart illustrating the treatment procedure and the number of patients who participated in this study.

Traditional patching control group (Patching Group): Patients were instructed to wear a standard, latex-free and adhesive style patch in front of their normal eyes for two hours, 7 days per week throughout the 12-week treatment period.

For all of the patients in Flicker Group and Patching Group, the flicker and patching treatment was combined with optimal refractive correction. The parents of the patients were given a form by which they reported a completion of the patching session each day throughout the treatment.

During the period of wearing the EFG or the patch, patients performed daily activities such as doing homework or watching television. During their first visit (Day 1), guardians of the patients within the Flicker Group were informed about the specifics of the EFG device, such as recharging and handling. Follow-up visits were scheduled after 3, 6 and 12 weeks from their first visit. During each visit, monocular and binocular visual functions were assessed. We had decided to follow up patients for 12 weeks because the investigators that first reported the benefit of the EFG therapy also followed up their patients for 12 weeks.

#### Visual acuit

Best-corrected visual acuity (BVCA) were measured separately for each eye using a national standard visual acuity chart according to the criteria for visual acuity test established by the National Health Commission of the People’s Republic of China (Logarithmic Visual Acuity Chart, xk100-06, CHINA). The total score within each line from the Logarithmic Visual Acuity Chart was 0.1 log units. Since there were five letters per line, the correctly read letter was assigned a score of 0.02 log units. The patients were tested with one eye at 5 m from the chart and the non-tested eye occluded throughout the test.

#### Contrast sensitivity

A CSV-1000 grating chart (VectorVision^®^ Inc., Greenville, OH 45331, USA) was used to measure the contrast sensitivity of the patients. This grating chart enabled us to test sensitivity for four spatial frequencies: 3, 6, 12 and 18 cycles per degree (cpd). This chart has been shown to be clinically reliable for tracking changes in visual performance of clinical patients [33]. We instructed the patients, who were equipped with optimal spectacle correction, to stand at a viewing distance of 40 cm in a dark room throughout the test.

#### Stereoacuity

Stereoacuity was assessed at a viewing distance of 40 cm under natural light with a Titmus Stereo test (Stereo Optical Co., Chicago, IL, USA). All patients wore polarizing glasses and, if necessary, additional optical corrections throughout testing. The Titmus Stereo test has been widely used in the clinical setting for measuring stereoacuity [34, 35]. If the patients were not able to perceive the largest disparity given by the Titmus Stereo test, we recorded that the stereoacuity of the patients was ‘nil’ and converted the non-numerical value into 4500 arc seconds for data analysis.

#### Fusion rang

A good fusion range can stabilize binocular vision. As an index of binocular vision, we measured fusion range between both eyes in all patients using Synoptophore L-2510B/L-2510HB (Inami & Co., Ltd., Japan). The scales in the device for measuring fusion range are degrees and Prism Dioptres (Δ).

### 2.4 Data analysis

Statistical analyses were performed using R software [36]. Data were visualized using Python’s matplotlib [37] and R’s ggplot2 [38] packages. A Shapiro-Wilk test indicated that our dataset did not assume a normal distribution (p < 0.05) and failed the meet the requirement for the use of parametric procedures. Therefore, we performed nonparametric (rank-based) analysis of variance (ANOVA)-like computation of longitudinal data using the package ‘nparLD’ designed for R software [39]. It is a rank-based finite sample approximation based on the quantiles from F-distribution. Post-hoc tests were performed with Bonferroni correction when there was either a significant main effect of treatment group or time. We report ANOVA-type statistics [40]. Effect sizes are reported as Cohen’s d. 95% Confidence intervals (CI) were obtained from t-distribution approximations.

## 3 Results

### 3.1 Baseline characteristics

We wanted to ensure that all patients were randomly assigned to the two treatment groups. So, by conducting statistical tests, we looked for possible differences between the Flickering and Patching groups based on the baseline visual performance. To do so, we used the nonparametric Wilcoxon-Mann-Whitney test. We found that the visual performance did not differ between the treatment groups (p-values ranged from 0.35 to 0.96) and concluded that the group assignment was random.

### 3.2 Compliance

We computed compliance by dividing the number of actual days where the patients had undergone the treatment at home from the number of total days for treatment. The mean compliance rate in the Flicker Group was 93.80 ± 0.025%, whereas the mean compliance rate in the Patching Group was 93.97 ± 0.021%. A nonparametric Wilcoxon-Mann-Whitney test revealed no significant difference in compliance rate between the two treatment groups (W = 116, p = 0.89).

### 3.3 Comparison between the EFG therapy and the standard patching therapy

Our study attempts to clarify whether the EFG therapy (i.e., Flicker Group) could be a robust and reliable treatment that could perhaps replace the standard patching therapy (i.e., Patching Group). To upend the standard treatment, it would first have to show whether the patients who had undergone the EFG therapy would show improvements in monocular and binocular visual functions. Second, it would have to bring about a significantly larger improvement than that from the standard patching therapy. Detailed statistical analyses are outlined below and revolve around these two central issues.

#### 3.3.1 Visual acuity (primary outcome)

We firstly conducted a nonparametric mixed ANOVA-like test (see Methods), which includes between-subject factor (treatment group) and within-subject factor (time). It revealed no main effect of group (F_(1.00,∞)_ = 1.57, p = 0.21) but a significant effect of time (F_(2.37,∞)_ = 40.42, p « 0.001). Then, to do post-hoc analysis, we performed pairwise multiple comparisons (with Bonferroni correction) for a significant main effect of within-subject factor time for each treatment group to determine if the primary outcome (defined as visual acuity at 12 weeks) significantly improved relative to baseline. There was a significant effect of within-subject factor time in the Flicker Group (p < 0.001, Cohen’s d = 0.86, 95% CI: [0.016, 0.24] logMAR) and in the Patching Group (p « 0.001, Cohen’s d = 1.08, 95% CI: [0.068, 0.34] logMAR). Individual data are plotted in Figure 3.

**Figure 3:**
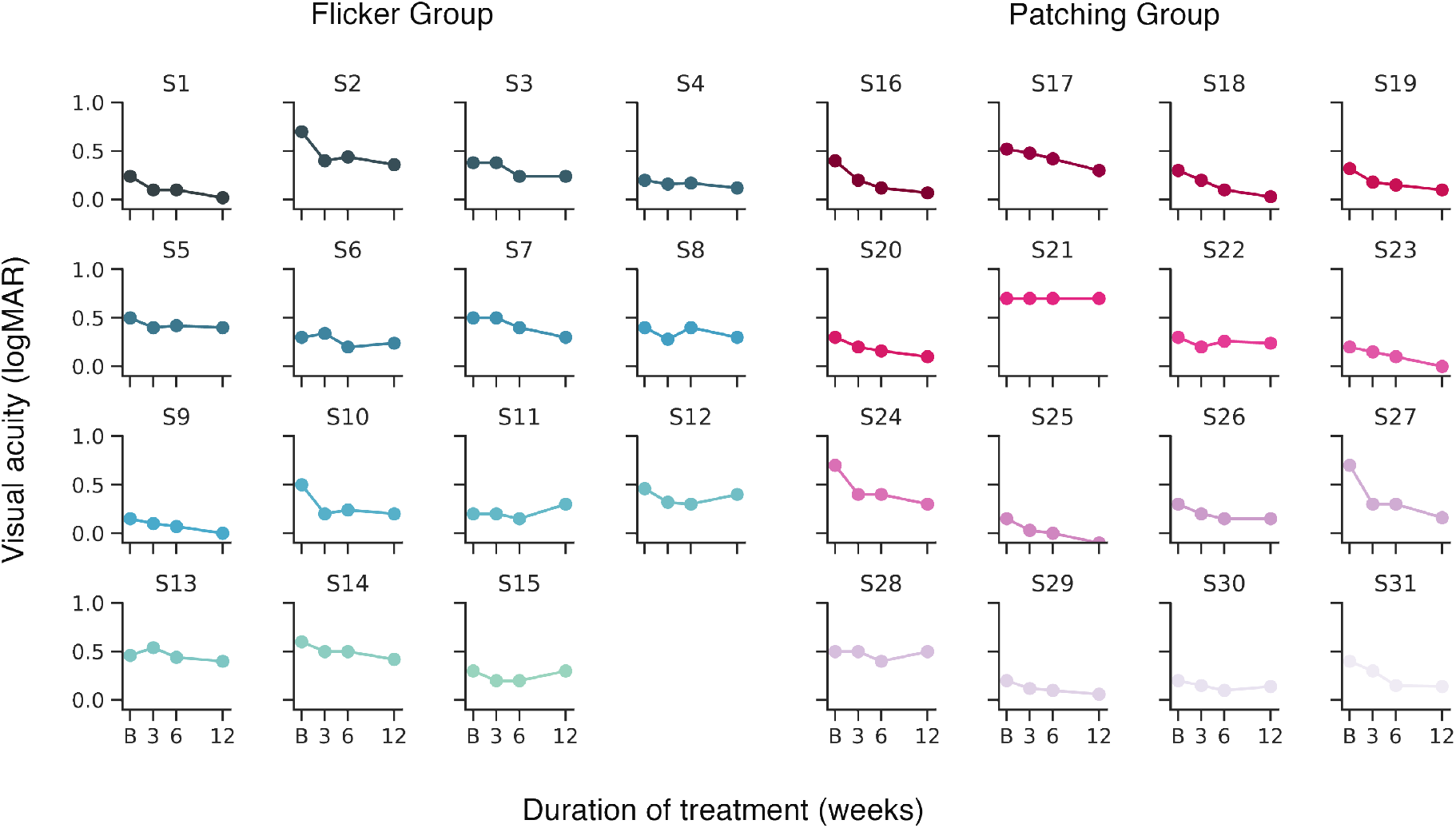
Visual acuity of each patient’s amblyopic eye at baseline and after beginning the treatment. ‘B’ denotes baseline measurement. Y-axis represents logMAR visual acuity, x-axis represent the time of measurement (in weeks) throughout the study.

#### 3.3 Comparison between the EFG therapy and the standard patching therapy

We also compared the improvement (Δ BVCA) between baseline and the three timepoints between the two groups by normalizing the visual acuity scores relative to baseline. The mixed ANOVA-like test (between-subject factor = treatment group, within-subject factor = time) revealed no main effect of group (F_(1.00,∞)_ = 2.26, p = 0.13) but a significant effect of time (F_(1.83,∞)_ = 9.93, p < 0.001). For post-hoc, pairwise multiple comparison was conducted (with Bonferroni correction) between the improvement in the Flicker Group and the Patching Group showed no difference (F_(1.00,∞)_ = 2.53, p = 0.11).

Also, we found a significant interaction between the treatment group and time (F_(2.29,∞)_ = 4.39, p = 0.0091). This interaction effect is illustrated in Figure 4A, which shows non-parallel time curves of logMAR visual acuity score in Flicker Group and Patching Group. The non-parallel plots suggest that the course of improvements over time in visual acuity significantly differed between the two treatment groups. According to our statistics and data (see Figure 4A), it seems that the rate of visual acuity improvement was significantly faster in the Patching Group than in the Flicker Group.

**Figure 4:**
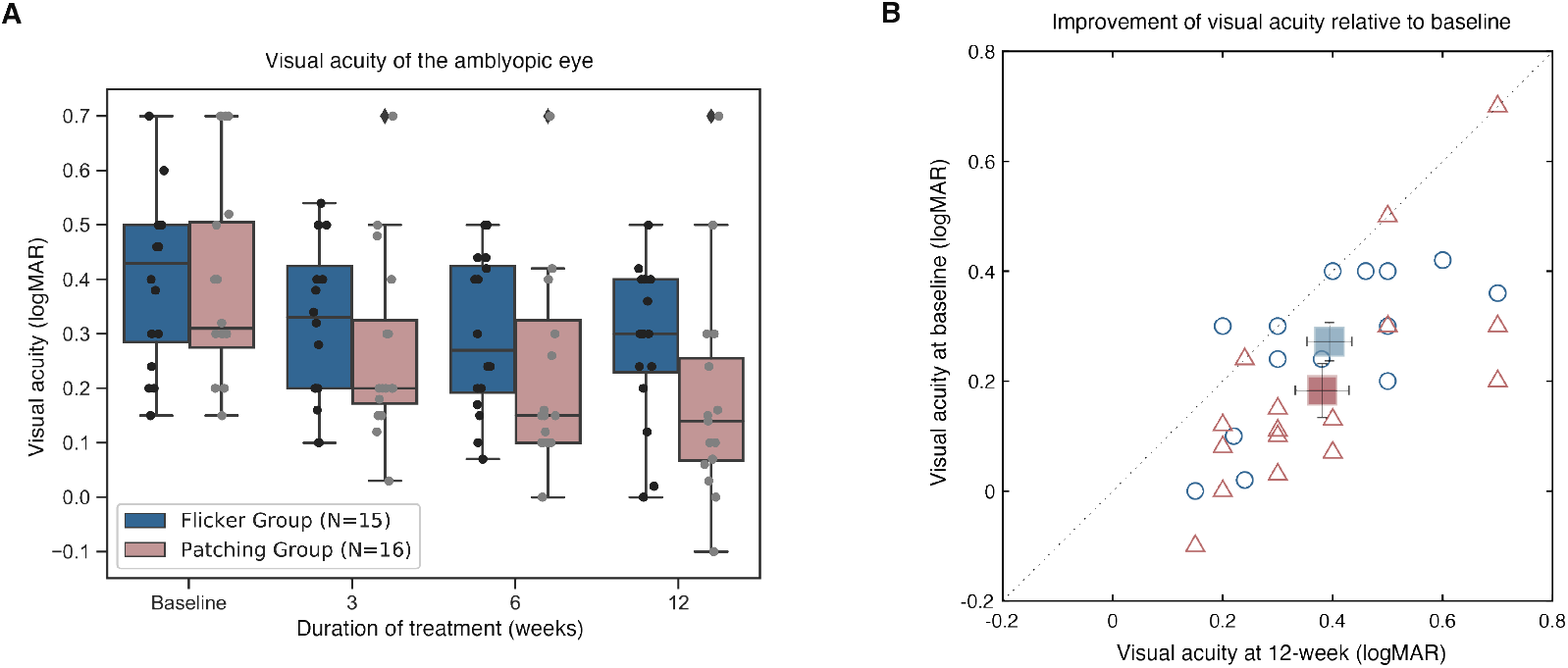
Best-corrected visual acuity. A) Boxplots of logMAR visual acuity in Flicker and Patching treatment groups. Blue represents the Flicker Group, pink the Patching Group. Individual data point is represented by a dot. The black solid line within each box represents the median. The box represents the interquartile range (IQR) of the data (25th to the 75th percentile). The whisker represents 1.5 × IQR either above the third quartile or below the first quartile. Diamonds represent outliers. B) Individual data point of visual acuity (BVCA) in patients’ amblyopic eyes measured at baseline and at 12 weeks of treatment. The dashed line represents unity. Blue circles represent the Flicker Group, whereas pink triangles represent the Patching Group. All points except six (three in the Flicker Group, three in the Patching Group) reside in below the dashed line, suggesting a notable improvement in visual acuity after 12 weeks of treatment. Any point that lies in the dashed line indicates no improvement after the therapies.

Except for three patients in the Flicker Group and another three in the Patching Group (points above the dashed line in Figure 4B), all experienced an improvement in visual acuity at 12 weeks relative to baseline (points below the dashed line in Figure 4B).

#### 3.3.2 Contrast sensitivity

We performed the non-parametric ANOVA-like test, which includes one between-subject factor (treatment group) and one within-subject factor (time) for every spatial frequency (3, 6, 12 and 18 cpd). Therefore, spatial frequency was not included as a within-subject factor for analysis. First, we found no main effect of treatment group from the non-parametric ANOVA but a significant effect of time at all spatial frequencies (p’s « 0.001). For post-hoc analysis, we performed pairwise multiple comparisons (with Bonferroni correction) between baseline and week 12 for each treatment group and spatial frequency. Comparisons with other combinations of timepoints (ex. 3-week vs. 6-week) are not reported for brevity and their poor clinical importance. Data in the form of fitted contrast sensitivity curves are shown in Figure 5.

**Figure 5:**
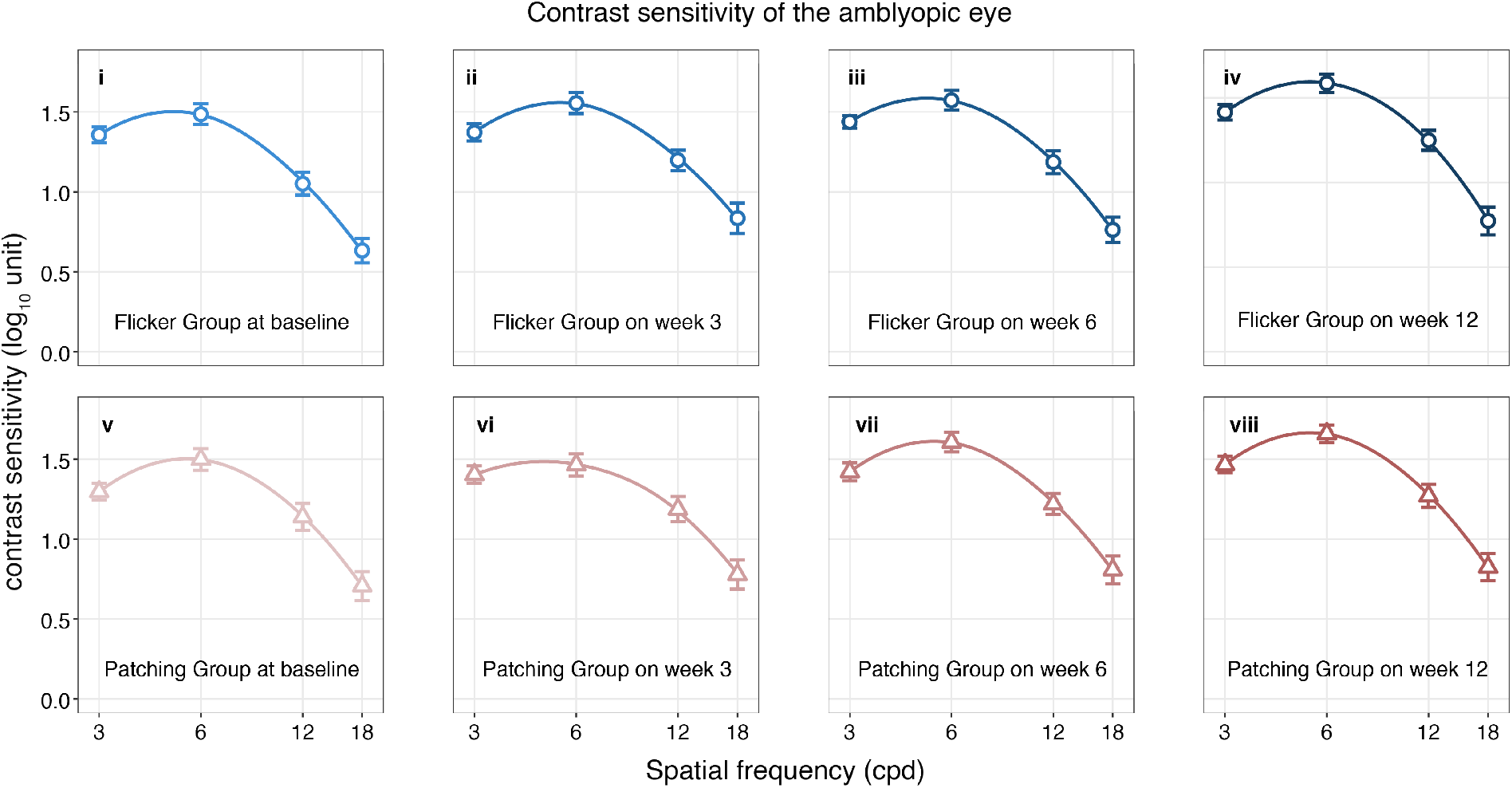
Means of contrast sensitivity for each treatment group at all time points of the treatment procedure. Blue plots represent the Flicker Group, pink plots the Patching Group. The points of circle represent the mean contrast sensitivity across patients in the Flicker Group, and the points of triangle the mean contrast sensitivity across patients in the Patching Group. The increased darkness for each color represents the increased period of treatment.

At 3 cpd, a pairwise comparison showed a significant difference in contrast sensitivity between baseline and week 12 in the Flicker Group (F_(1,∞)_ = 8.70, p = 0.0031; Cohen’s d = 0.57) and the Patching Group (F_(1,∞)_ = 6.82, p = 0.027; Cohen’s d = 0.94). At 6 cpd, it showed no significant difference between the timepoints in the Flicker Group (F_(1,∞)_ = 4.98, p < 0.001; Cohen’s d = 0.77) but did so in the Patching Group (F_(1,∞)_ = 17.0, p < 0.001; Cohen’s d = 1.06). In the Flicker Group the null hypothesis was not rejected; however, we cannot simply accept the alternative hypothesis because the effect size is large (> 0.5). It could be that we had an inadequate sample size since it had been established based on the population data of visual acuity (see Methods), which was our primary outcome. At 12 cpd, it showed a significant difference in the Flicker Group (F_(1,∞)_ = 8.70, p = 0.0095; Cohen’s d = 0.94) and in the Patching Group (F_(1,∞)_ = 6.50, p = 0.032; Cohen’s d = 0.98). At 18 cpd, it revealed no significant difference in the Flicker Group (F_(1,∞)_ = 2.93, p = 0.26; Cohen’s d = 0.38) and the Patching Group (F_(1,∞)_ = 3.59, p = 0.17; Cohen’s d = 0.75). The effect size was large (Cohen’s d > 0.5) in the Patching Group, but the null hypothesis was not rejected in the ANOVA test; this again could be attributed to the sample size. Therefore, alternative hypothesis could not be accepted with certainty.

#### 3.3.3 Stereopsis

Individual data are provided in Figure 6. Nonparametric mixed ANOVA-like test revealed no main effect of group (F_(1,∞)_ = 0.52, p = 0.47) and no significant effect of time (F_(2.46,∞)_ = 2.18, p = 0.10). Also, it revealed a no interaction effect between the treatment group and time (F_(2.46,∞)_ = 0.56, p = 0.61). These statistical findings can be confirmed in Figure 7, which shows no difference between the groups at all timepoints.

**Figure 6:**
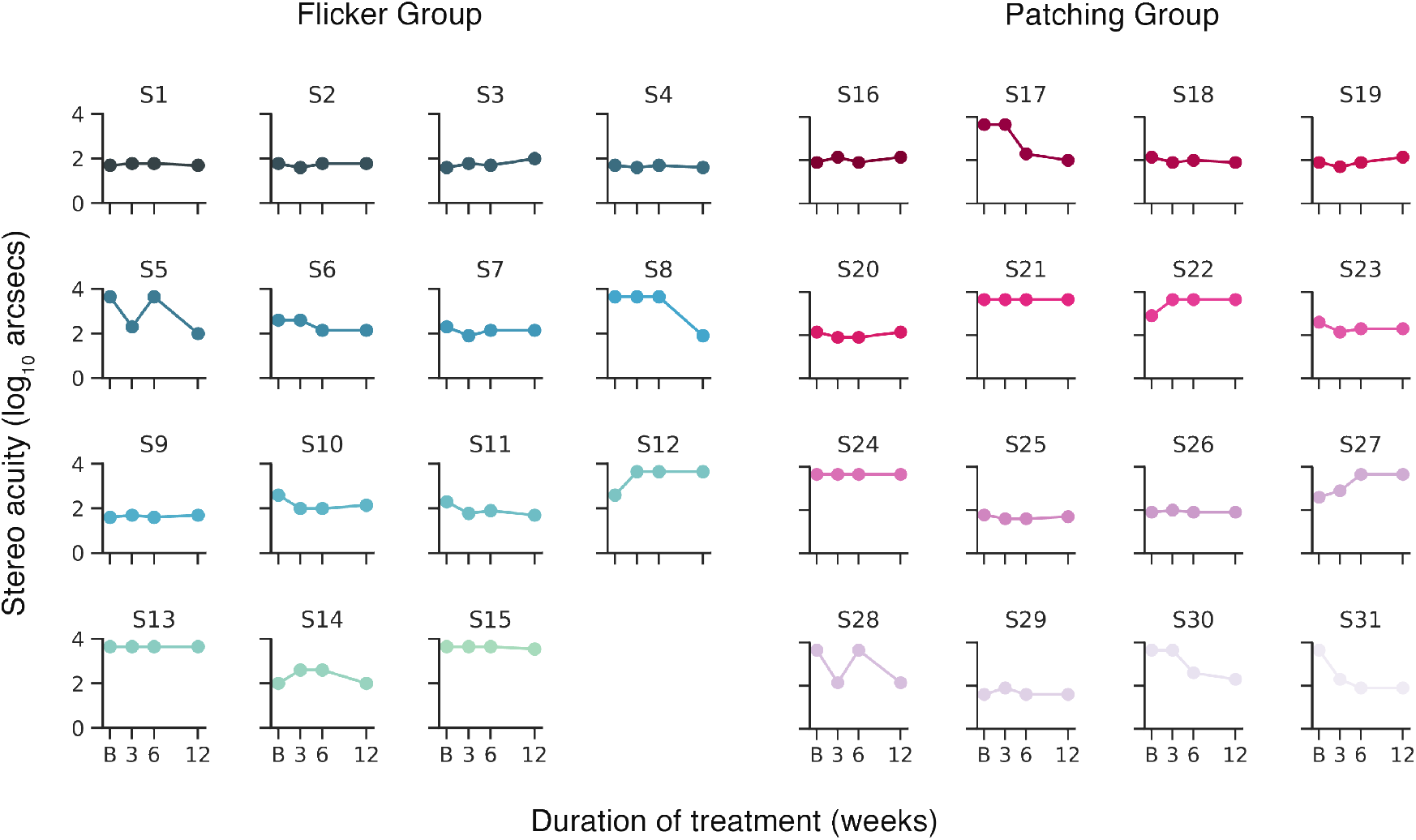
Individual patient’s stereoacuity at baseline and after beginning the treatment. ‘B’ denotes baseline measurement. Y-axis represents stereo threshold that has been log transformed (log10 of the original stereo threshold), x-axis represent the time of measurement throughout the study.

**Figure 7:**
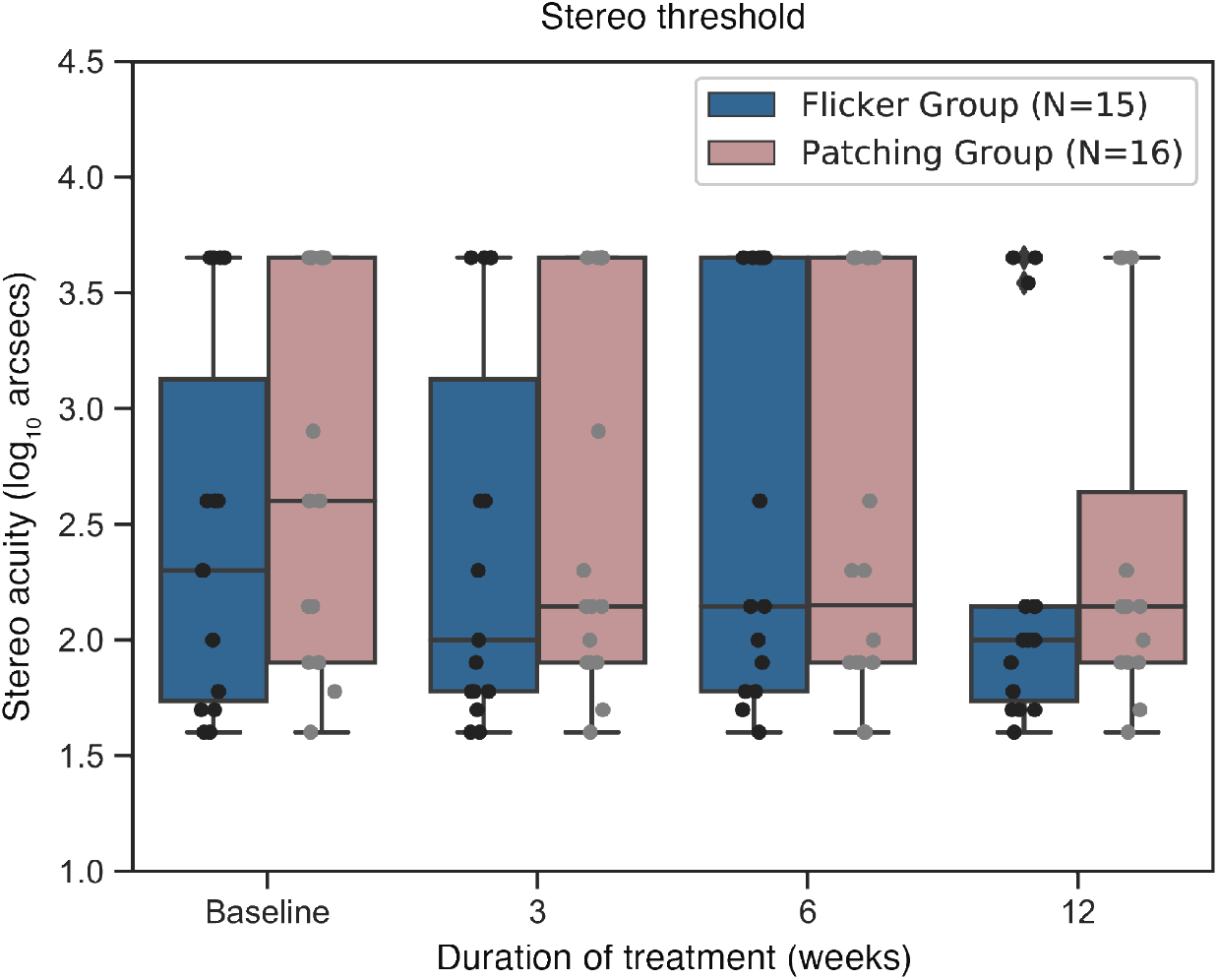
Boxplots of stereo thresholds in Flicker and Patching treatment groups. Blue represents the Flicker Group, pink the Patching Group. Individual data point is represented by a dot. The black solid line within each box represents the median. The box represents the interquartile range (IQR) of the data (25th to the 75th percentile). The whisker represents 1.5 × IQR either above the third quartile or below the first quartile. Diamonds represent outliers.

#### 3.3.4 Fusion range

Five patients (two in the Flicker Group, three in the Patching Group) were not able to complete the study and therefore had to be excluded in data analysis (see Figure 8). Nonparametric ANOVA-like test showed no main effect of group (F_(1,∞)_ = 0.98, p = 0.32) and a significant effect of time (F_(2.60,∞)_ = 3.27, p = 0.026). It revealed a no interaction effect between the treatment group and time (F_(2.60,∞)_ = 0.77, p = 0.50). Then, we performed post hoc tests for a significant main effect of within-subject factor time between baseline and week 12 for each treatment group. We observed a significant difference in fusion range between baseline and week 12 in the Flicker Group (p = 0.015, Cohen’s d = 0.41) but not in the Patching Group (p = 0.33, Cohen’s d = 0.33). The fact that the null hypothesis was rejected and that the effect size was large in the Flicker Group indicates that fusion range was improved more so in the Flicker Group throughout the study than in the Patching Group (see Figure 9).

**Figure 8:**
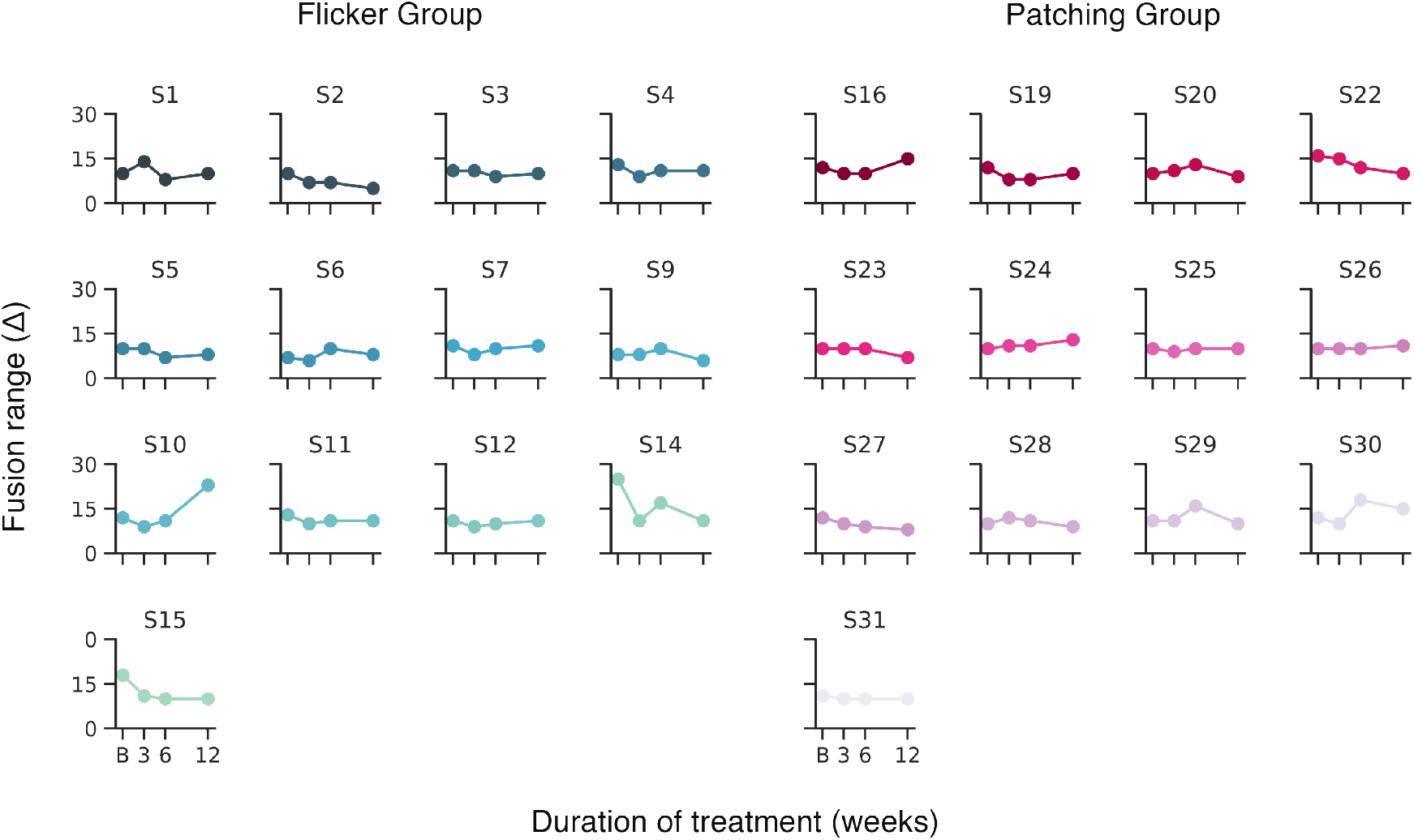
Individual patient’s fusion range at baseline and after beginning the treatment. ‘B’ denotes baseline measurement. Y-axis represents fusion range, x-axis represent the time of measurement throughout the study.

**Figure 9:**
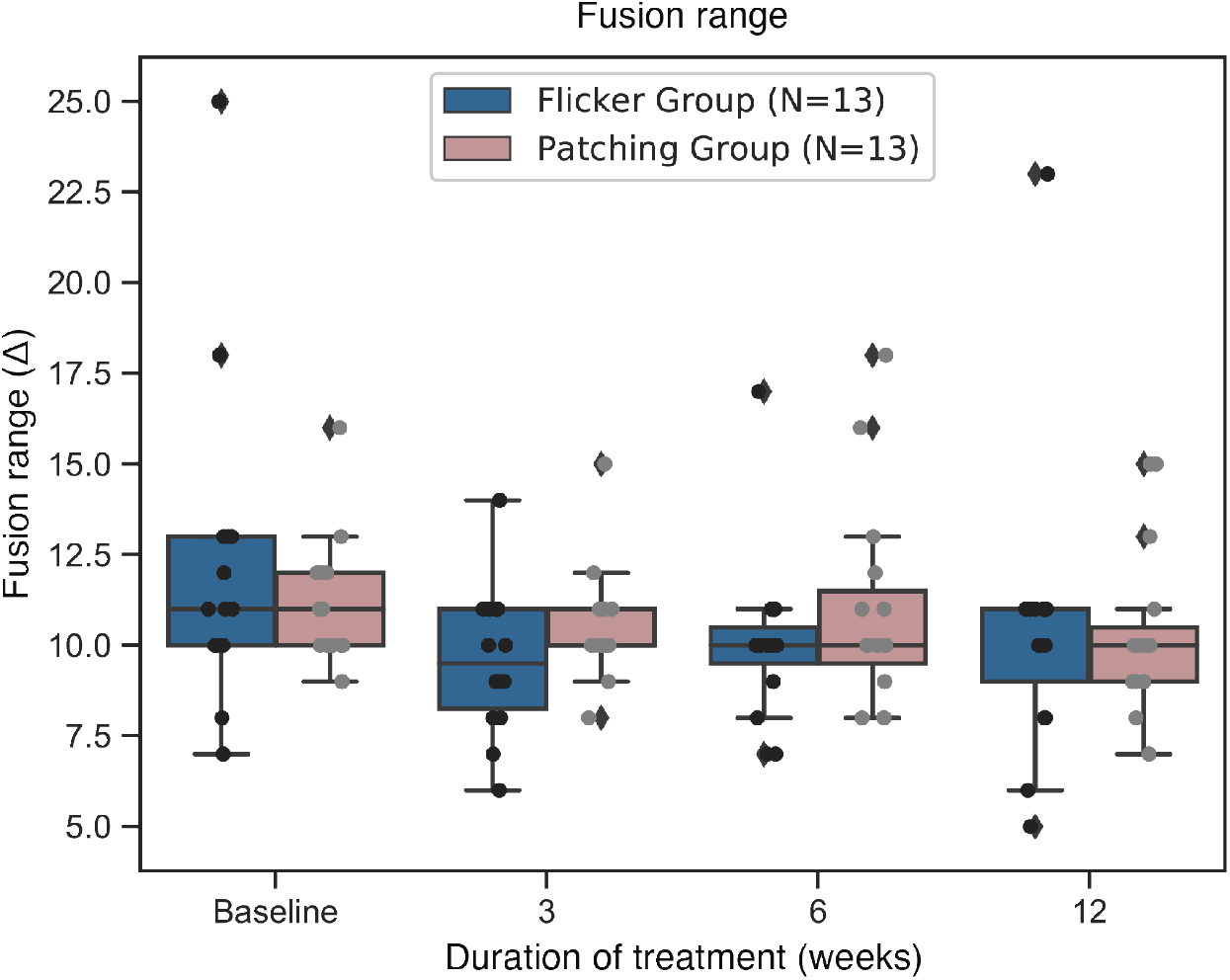
Boxplots of fusion range in Flicker and Patching treatment groups. Blue represents the Flicker Group, pink the Patching Group. Individual data point is represented by a dot. The black solid line within each box represents the median. The box represents the interquartile range (IQR) of the data (25th to the 75th percentile). The whisker represents 1.5 × IQR either above the third quartile or below the first quartile. Diamonds represent outliers.

## 4 Discussion

According to the literature, this study is the first randomized controlled trial that compares the efficacy of Eyetronix Flicker Glass and adhesive occlusion patches in children with amblyopia. We measured amblyopic eye’s visual acuity (BVCA), its contrast sensitivity to measure monocular functions and stereopsis and fusion range to measure binocular functions.

As for visual acuity at 12 weeks, which was our primary outcome for this study, we observed that both treatment groups (Flicker and Patching) resulted in a significant improvement in visual acuity at 12 weeks relative to baseline. The rate of improvement, however, was more rapid in the Patching Group. For contrast sensitivity, both treatment groups experienced a significant improvement at 3, 6 and 12 cpd (except at 6cpd for the Flicker Group) but not at 18 cpd. This suggests that at high spatial frequency where amblyopes experience poor perception, the improvement was smaller than those at low spatial frequency. On the other hand, stereopsis did not improve in both treatment groups. As for fusion range, the Flicker Group experienced a significant improvement at 12 weeks relative to baseline but not the Patching Group.

The ranges of improvement in visual acuity (from 95% CI and effect sizes) in the two treatment groups seem to be similar (see Results). This is surprising given the fact that the deprivation period for the normal eye in the Flicker Group was equivalent to only 30 minutes (50% of daily deprivation due to alternate flicker), whereas the one in the Patching Group was 2 hours. This indicates that duration of deprivation might not be important. In other words, longer deprivation might not be linearly beneficial. If this holds true, then the compliance rate of amblyopic children could be higher relative to that observed during the standard patching therapy. However, the rate of improvement in visual acuity was significantly different between the two groups; the Patching Group improved faster.

Patients in the Flicker Group experienced an improvement in fusion range, which is a binocular function, at 12 weeks of treatment relative to baseline. As previously mentioned, EFG therapy has been introduced on the premise that alternating visual flicker can reduce suppression and improve binocular vision [30]. Our findings are in line with the premise. Our study therefore advocates the notion that binocular treatment strategy, which manipulates visual input in both eyes, brings about binocular benefits for amblyopic patients. However, our findings did not show improvements in stereopsis, as it was reported in a previous study (Vera-Diaz et al., 2016) [32]. This can be due to multiple reasons. 1) Occlusion period was shorter in our study. The two previous studies employed 1-2 hours of occlusion as opposed to a fixed number of hours like in our study. 2) The age distribution of the patients seems to be notably different between our study and the previous study. For instance, all fifteen patients (in Flicker Group) from our study were aged four to nine years old. On the other hand, twelve out of twenty-three patients in the study of Vera-Diaz et al. (2016) were above ten years old. However, despite the different age distributions in the patients across the studies, all of them had anisometropic amblyopia. In future, shorter and longer treatment durations of the EFG therapy could also be explored in future studies with the hope to find the optimal duration of the therapy.

## 5 Limitations

Our study has several limitations. First, since all of our patients were anisometropic, the implications of our findings might not be generalizable to other types of amblyopia, such as strabismic and deprivation amblyopia. Second, we did not equally deprive the normal eye for each treatment group. The patients in the Patching Group underwent 2-hour deprivation of the fellow eye and those in the Flicker Group 30-minute deprivation of the fellow eye (in a 1-hour treatment). Third, we did not perform follow-up assessments of the patients after the end of the treatment. Therefore, whether the monocular benefit from the EFG therapy would last remains to be investigated. Lastly, our sample size had been established based on the population standard deviation of visual acuity. Therefore, although we can confirm that its power was adequate for us to reject the null hypothesis, we cannot do so for the some of our spatial frequency data.

## 6 Conclusion

In children aged 4-13 years old with moderate or several unilateral anisometropic amblyopia, both the standard patching therapy (i.e., patching the fellow eye for two hours/day) and the EFG therapy (i.e., alternative flicker for one hour/day) were effective in improving visual acuity of the amblyopic eye. The two therapies are not different in terms of the visual acuity improvement at 12-week of the amblyopic eye. Both therapies did not improve stereopsis after 12 weeks of treatment. Contrast sensitivity at 3, 6 and 12 cpd was improved in both groups but not so at 18 cpd. Fusion range was significantly improved in the Flicker Group but not in the Patching Group. Therefore, our study suggests that the EFG therapy resulted in both monocular and binocular benefits for amblyopic patients, whereas the traditional patching therapy only brought about monocular benefits.

## Data Availability

Data supporting the findings of this study are available from the corresponding author upon request.

## 7 Acknowledgment

This research was supported by National Natural Science Foundation of China (Grant Nos. 31970975 and 81500754) and the Wenzhou Medical University grant (QTJ16005) to JWZ and Canadian institute of Health Research’s graduate award to SHM.

## References

[1] Jonathan M Holmes and Michael P Clarke. Amblyopia. The Lancet, 367(9519):1343–1351, 2006.

[2] DM Levi and RS Harwerth. Contrast evoked potentials in strabismic and anisometropic amblyopia. Investigative Ophthalmology & Visual Science, 17(6):571–575, 1978.

[3] A Bradley and RD Freeman. Contrast sensitivity in anisometropic amblyopia. Investigative ophthalmology & visual science, 21(3):467–476, 1981.

[4] RF Hess and ER Howell.The threshold contrast sensitivity function in strabismic amblyopia: evidence for a two type classification. Vision research, 17(9):1049–1055, 1977.

[5] Jan Walraven and Piet Janzen. Tno stereopsis test as an aid to the prevention of amblyopia. Ophthalmic and Physiological Optics, 13(4):350–356, 1993.

[6] R Thad Goodwin and Paul E Romano. Stereoacuity degradation by experimental and real monocular and binocular amblyopia. Investigative ophthalmology & visual science, 26(7):917–923, 1985.

[7] Robert F Hess and Benjamin Thompson. Amblyopia and the binocular approach to its therapy. Vision research, 114:4–16, 2015.

[8] Dennis M Levi, David C Knill, and Daphne Bavelier. Stereopsis and amblyopia: A mini-review. Vision research, 114:17–30, 2015.

[9] C De Buffon. Disseration sur la cause du strabisme ou des yeux louches. Me. Acad Roy Sci, 1743, 1743.

[10] Michael X Repka, Roy W Beck, Jonathan M Holmes, Eileen E Birch, Danielle L Chandler, Susan A Cotter, Richard W Hertle, Raymond T Kraker, Pamela S Moke, Graham E Quinn, et al. A randomized trial of patching regimens for treatment of moderate amblyopia in children. Archives of ophthalmology (Chicago, Ill.: 1960), 121(5):603–611, 2003.

[11] Maria Fronius, Licia Cirina, Hanns Ackermann, Thomas Kohnen, and Corinna M Diehl. Efficiency of electronically monitored amblyopia treatment between 5 and 16 years of age: new insight into declining susceptibility of the visual system. Vision research, 103:11–19, 2014.

[12] Catherine E Stewart, Merrick J Moseley, David A Stephens, and Alistair R Fielder. Treatment dose-response in amblyopia therapy: the monitored occlusion treatment of amblyopia study (motas). Investigative ophthalmology & visual science, 45(9):3048–3054, 2004.

[13] Michael P Wallace, Catherine E Stewart, Merrick J Moseley, David A Stephens, and Alistair R Fielder. Compliance with occlusion therapy for childhood amblyopia. Investigative ophthalmology & visual science, 54(9):6158–6166, 2013.

[14] A Searle, P Norman, R Harrad, and K Vedhara. Psychosocial and clinical determinants of compliance with occlusion therapy for amblyopic children. Eye, 16(2):150–155, 2002.

[15] Eileen E Birch. Amblyopia and binocular vision. Progress in retinal and eye research, 33:67–84, 2013.

[16] Yiya Chen, Zhifen He, Yu Mao, Hao Chen, Jiawei Zhou, and Robert F Hess. Patching and suppression in amblyopia: one mechanism or two? Frontiers in Neuroscience, 13, 2019.

[17] Eileen E Birch, Yolanda S Castañeda, Christina S Cheng-Patel, Sarah E Morale, Krista R Kelly, Cynthia L Beauchamp, and Ann Webber. Self-perception of school-aged children with amblyopia and its association with reading speed and motor skills. JAMA ophthalmology, 137(2):167–174, 2019.

[18] Stephen B Kaye, Sean I Chen, Gary Price, Lesley C Kaye, Carmel Noonan, Ajay Tripathi, Pammal Ashwin, Nolan Cota, David Clark, and Jeremy Butcher. Combined optical and atropine penalization for the treatment of strabismic and anisometropic amblyopia. Journal of American Association for Pediatric Ophthalmology and Strabismus, 6(5):289–293, 2002.

[19] Omry BenEzra, Rafi Herzog, Evelyne Cohen, Ilana Karshai, and David BenEzra. Liquid crystal glasses: feasibility and safety of a new modality for treating amblyopia. Archives of ophthalmology, 125(4):580–581, 2007.

[20] Omry BenEzra, Abraham Spierer, and Oriel Spierer. Liquid crystals—transformer that meets the eye. Journal of pediatric ophthalmology and strabismus, 52(5):266–267, 2015.

[21] Ibrahim Erbağci, Seydi Okumuş, Veysi Öner, Erol Coşkun, Oğuz Çelik, and Burak Ören. Using liquid crystal glasses to treat amblyopia in children. Journal of American Association for Pediatric Ophthalmology and Strabismus, 19(3):257–259, 2015.

[22] Abraham Spierer, Judith Raz, Omry BenEzra, Rafi Herzog, Evelyne Cohen, Ilana Karshai, and David BenEzra. Treating amblyopia with liquid crystal glasses: a pilot study. Investigative ophthalmology & visual science, 51(7):3395–3398, 2010.

[23] Daniel H Baker, Tim S Meese, and Robert F Hess. Contrast masking in strabismic amblyopia: attenuation, noise, interocular suppression and binocular summation. Vision research, 48(15):1625–1640, 2008.

[24] Jian Ding, Stanley A Klein, and Dennis M Levi. Binocular combination in abnormal binocular vision. Journal of vision, 13(2):14–14, 2013.

[25] Karen Holopigian, Randolph Blake, and Mark J Greenwald. Selective losses in binocular vision in anisometropic amblyopes. Vision research, 26(4):621–630, 1986.

[26] Chang-Bing Huang, Jiawei Zhou, Zhong-Lin Lu, and Yifeng Zhou. Deficient binocular combination reveals mechanisms of anisometropic amblyopia: Signal attenuation and interocular inhibition. Journal of vision, 11(6):4–4, 2011.

[27] MiYoung Kwon, Emily Wiecek, Steven C Dakin, and Peter J Bex. Spatial-frequency dependent binocular imbalance in amblyopia. Scientific reports, 5:17181, 2015.

[28] Jiawei Zhou, Alexandre Reynaud, Zhimo Yao, Rong Liu, Lixia Feng, Yifeng Zhou, and Robert F Hess. Amblyopic suppression: Passive attenuation, enhanced dichoptic masking by the fellow eye or reduced dichoptic masking by the amblyopic eye? Investigative ophthalmology & visual science, 59(10):4190–4197, 2018.

[29] Pi-Chun Huang, Daniel H Baker, and Robert F Hess. Interocular suppression in normal and amblyopic vision: spatio-temporal properties. Journal of vision, 12(11):29–29, 2012.

[30] Clifton Schor, Mark Terrell, and Donald Peterson. Contour interaction and temporal masking in strabismus and amblyopia. American journal of optometry and physiological optics, 53(5):217–223, 1976.

[31] Jingyun Wang, Daniel E Neely, Jay Galli, Joshua Schliesser, April Graves, Tina G Damarjian, Jessica Kovarik, James Bowsher, Heather A Smith, Dana Donaldson, et al. A pilot randomized clinical trial of intermittent occlusion therapy liquid crystal glasses versus traditional patching for treatment of moderate unilateral amblyopia. Journal of American Association for Pediatric Ophthalmology and Strabismus, 20(4):326–331, 2016.

[32] Moore B. Hussey E. Srinivasan G. Johnson C. Vera-Diaz, F. A. A flicker therapy for the treatment of amblyopia. Vision Development and Rehabilitation, 2(2):105–114, 2016.

[33] Glenn N Pomerance and David W Evans. Test-retest reliability of the csv-1000 contrast test and its relationship to glaucoma therapy. Investigative ophthalmology & visual science, 35(9):3357–3361, 1994.

[34] Elise B Ciner, Gui-shuang Ying, Marjean Taylor Kulp, Maureen G Maguire, Graham E Quinn, Deborah Orel-Bixler, Lynn A Cyert, Bruce Moore, and Jiayan Huang. Stereoacuity of preschool children with and without vision disorders. Optometry and vision science: official publication of the American Academy of Optometry, 91(3):351, 2014.

[35] David K Wallace, Elizabeth L Lazar, Michele Melia, Eileen E Birch, Jonathan M Holmes, Kristine B Hopkins, Raymond T Kraker, Marjean T Kulp, Yi Pang, Michael X Repka, et al. Stereoacuity in children with anisometropic amblyopia. Journal of American Association for Pediatric Ophthalmology and Strabismus, 15(5):455–461, 2011.

[36] J Allaire. Rstudio: integrated development environment for r. Boston, MA, 770:394, 2012.

[37] John D Hunter. Matplotlib: A 2d graphics environment. Computing in science & engineering, 9(3):90–95, 2007.

[38] Hadley Wickham. ggplot2: elegant graphics for data analysis. springer, 2016.

[39] Kimihiro Noguchi, Yulia R Gel, Edgar Brunner, and Frank Konietschke. nparld: an r software package for the nonparametric analysis of longitudinal data in factorial experiments. Journal of Statistical Software, 50(12), 2012.

[40] Edgar Brunner, Frank Konietschke, Markus Pauly, and Madan L Puri. Rank-based procedures in factorial designs: hypotheses about non-parametric treatment effects series b statistical methodology. 2017.

